# Summarizing Clinical Notes using LLMs for ICU Bounceback and Length-of-Stay Prediction

**DOI:** 10.1101/2025.01.19.25320797

**Authors:** Akash Choudhuri, Philip Polgreen, Alberto Segre, Bijaya Adhikari

## Abstract

Recent advances in the Large Language Models (LLMs) provide a promising avenue for retrieving relevant information from clinical notes for accurate risk estimation of adverse patient outcomes. In this empirical study, we quantify the gain in predictive performance obtained by prompting LLMs to study the clinical notes and summarize potential risks for downstream tasks. Specifically, we prompt LLMs to generate a summary of progress notes and state potential complications that may arise. We then learn representations of the generated notes in sequential order and estimate the risks of patients in the ICU getting readmitted in ICU after discharge (ICU bouncebacks) and predict the overall length of stay in the ICU. Our analysis in the real-world MIMIC III dataset shows performance gains of 7.17% in terms of AUC-ROC and 14.16% in terms of AUPRC for the ICU bounceback task and 2.84% in terms of F-1 score and 7.12% in terms of AUPRC for the ICU LOS Prediction task. This demonstrates that the LLM-infused models outperform the approaches that only directly rely on clinical notes and other EHR data.

## I. Introduction

Estimating the risk of an inpatient’s condition worsening is crucial in healthcare facilities, as the identification of high-risk patients aids in strategic hospital decision-making [1], and the application of proactive preventive measures enables early intervention. The fine-grained information on patients’ trajectories embedded within Electronic Healthcare Records (EHRs) makes patient risk estimation feasible. Recent advances in machine learning have brought significant strides in EHR analytics; specific examples include extraction of patient risk factors [2]–[4], leveraging the underlying data storage structures of EHRs for representation learning [5]–[8], and the inference of interactions between healthcare entities [9], [10]. This line of research has produced scalable and highly accurate frameworks for patient risk estimation in healthcare facilities [11].

Despite advances in machine learning, most prior works in this space fail to effectively capture the rich information stored in unstructured free-text clinical notes. Clinical notes contain subtle spectra of individual patient risk factors that reflect the direct perspective of physicians and healthcare workers and are not necessarily captured by tabular records. There has been some recent interest [5], [10] in mining clinical notes along with other data sources for downstream predictive tasks. However, these approaches learn representations only from the text present in the clinical notes and fail to capture the knowledge that exists outside clinical nodes, for e.g. those in PubMed [12] and public forums like reddit [13]. The absence of this information poses a detrimental effect on effective knowledge mining. Although additional guidance can be externally provided via knowledge graphs (KGs) [14], [15], such a procedure requires caution in aligning the concepts to their corresponding meaning in the given EHR data as concepts and their meanings evolve over time [16].

Recent advances in Large Language Models (LLMs) in the domain of healthcare analytics [17]–[19] provide a promising way to resolve these issues, as they contain billions of parameters and have been pre-trained on massive corpora including text data from PubMed and public forums, thus inherently capturing a significant amount of external knowledge. Recent works like [20], [21] use LLMs on EHRs, but only work on hospital codes and fail to fully utilize the knowledge of LLMs and clinical notes simultaneously. However, LLMs enable retrieving the most meaningful information from clinical notes. To address this gap, our study empirically quantifies the degree of enhancement in the information obtained from clinical notes with LLMs to improve patient risk estimation. We hypothesize that the information obtained from LLMs fused with clinical notes provides more information than the clinical notes themselves, and we empirically show that the text generated by LLMs provides more evident risk factors that can aid in decision-making and allocation of resources in healthcare facilities. The contributions of our study are as follows:

- We quantitatively evaluate the integration of LLMs to clinical notes to enhance the information provided by clinical notes by providing potential medical complications that may occur in free text.
- We propose an end-to-end framework that integrates both tabular features and the sequential progression of risk in the form of textual data generated by LLMs for accurate patient risk estimation.
- We perform experiments on real-world and open-source EHR dataset MIMIC-III on two applications: ICU Bounceback Prediction and ICU Length of Stay Prediction tasks.

## II. Method

In this section, we will provide an overview of our methodology. The detailed overview of our overall framework is shown in Figure 1. Our methodology mainly consists of four steps, namely data extraction, large language model information extraction, temporal embedding of the generated summaries, and final prediction. We will first formulate the problem and then describe each component in detail.

**Figure 1:**
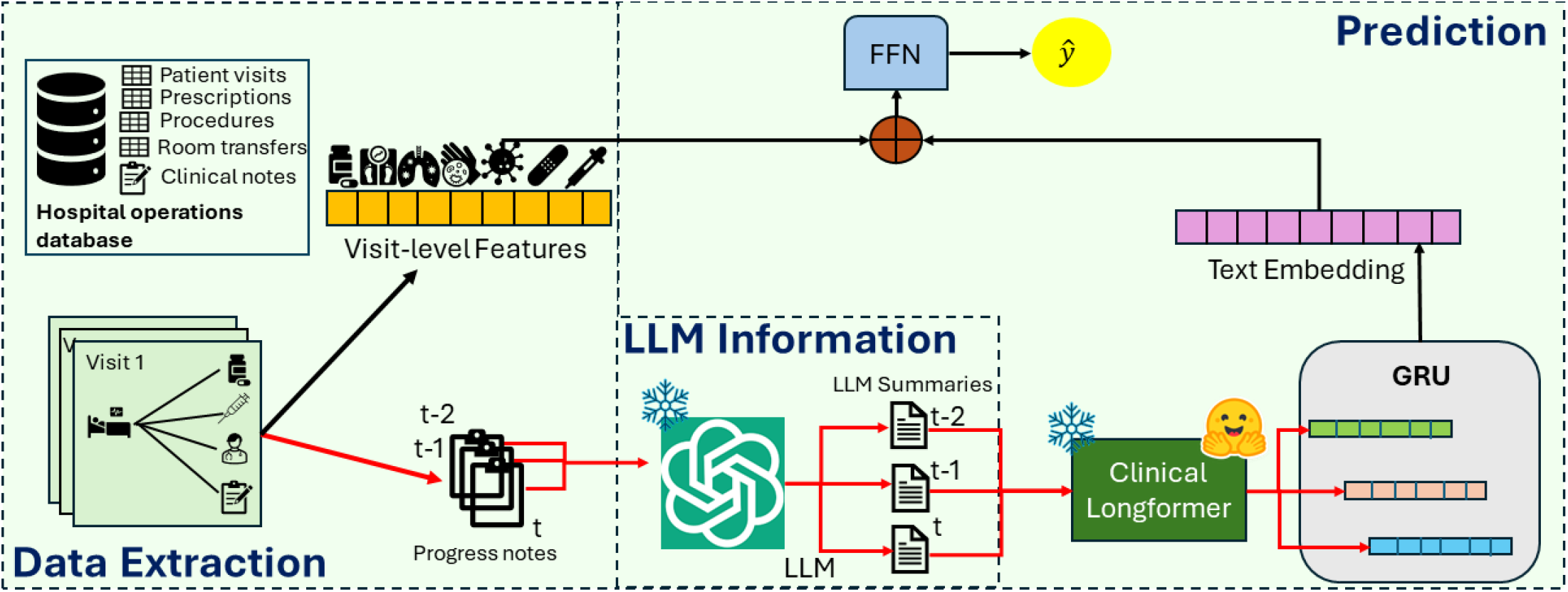
Proposed framework (best viewed in color). The steps denoted by red arrows are performed separately than the steps denoted by black arrows. Data Extraction constructs visit-level data and progress notes for each individual visit from the Hospital Operations Database. This data is then used to construct the visit-level features. The progress notes are sequentially inputted to the frozen LLM to generate summaries. Frozen Clinical Longformer generates embeddings of the corresponding summaries and these embeddings are sequentially passed through the GRU to generate the overall text embedding for the visit. This embedding is concatenated with the visit-level features and passed through the FFN to get the predictions.

### A. Problem Formulation

We are given a hospital operations database with events derived from EHRs [9], [10], [22], [23] and Admission Discharge Transfer (ADT) logs [9], [10], [24], [25] from an inpatient healthcare facility. The data contains time-stamped information about patient movement throughout the hospital as well as time-stamped records of procedures, laboratory tests, and prescribed medications. In addition to the items mentioned earlier, the data also contains time-stamped records of admission to critical-care units as well as unstructured clinical notes. This data can be used to extract information about each patient visit. The set of patient visits is denoted by 𝒱 Similarly, the corresponding patient activity data extracted from EHR and ADT databases can be denoted by 𝒳 _*i*_, where *i* ∈ 𝒱. Note that 𝒳 _*i*_ also contains clinical note data in addition to the other tabular data.

In addition to patient activity data, we are also given corresponding task labels *y*_*i*_ corresponding to each visit *i* ∈𝒱 Each task label indicates the eventual outcome that occurred after the patient’s visit. Examples include binary mortality labels, where positive labels could indicate the patient’s death after the current visit, and negative labels for otherwise. We can now formally define our problem.

**Given**

Patient visit activity data *{𝒳* _*i*_*}*_*i*∈ 𝒱_ for a set of patient visits 𝒱 and corresponding labels *{y*_*i*_*}* _*i*∈ 𝒱_

**Infer**

A mapping function *m*(.) which maps each visit data 𝒳 _*i*_ to corresponding label *y*^*i*^, where *i* ∈ 𝒱.

**Such that**

a loss function ∑_*i*∈𝒱_ *𝕃* (𝒳 _*i*_, *y*_*i*_) is minimized.

In the problem above, L is a standard classification loss function such as the cross-entropy loss. We solve this problem as a supervised classification problem, where each sample corresponds to a patient visit.

### B. Data Extraction

The data extraction module aims to leverage the relational structure of EHR data to extract relevant information required as inputs for the latter components of the framework. This step is used to extract both the visit-level as well as the unstructured clinical progress notes in the chronological order of entry into the system.

To extract the visit-level information, the database is queried to obtain the visit records and associated information relating to the corresponding visit (medications prescribed, procedures performed, possible diagnoses, etc.). This associated information will then be used to compute different comorbidity scores which are used as risk factors for patient health risk. On the other hand, demographic information like age, gender, race, etc is also extracted. This creates the tabular visit-level features *d*^*i*^ for every visit *i* ∈𝒱from 𝒳 _*i*_.

For unstructured clinical notes present in 𝒳_*i*_, we make sure to exclude discharge summaries from our data as they do not provide detailed information about the progress of the patient’s health status. Moreover, some discharge summaries could also mention the overall length of stay or the chances of readmission (our applications, which are described in Section IV.C.) and could thus lead to information leakage in our predictive task. So, for every patient visit *i* ∈ 𝒱, spanning from timestamp *T*_0_ to *T*_*T*_ we chronologically extract the progress notes denoted by 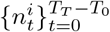 that dynamically document each patient’s health status. The exact details of extracting the progress notes for our experiments are given in Section IV.A.

### C. Large Language Model Information Extraction

Generative language models (GLMs) are advanced natural language processing models capable of producing text that is coherent and contextually relevant. Through extensive pretraining on large amounts of text data and fine-tuning based on human instructions, they can generate text outputs that closely resemble human-written content. LLMs model the probability of a sentence (that is, a sequence of word tokens)*s* = (*q*_1_, *q*_2_, …, *q*_*n*_) as 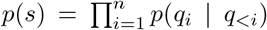, where *q*_*i*_ denotes the *i*-th token of the sentence *s* and *q*_*<i*_ denotes the partial word token sequence before the *i*-th step. Moreover, due to their training in wide corpora that encompasses multiple sources of information, LLMs have recently shown increased reasoning abilities in the medical domain [26]–[28] and have been deployed in various applications in conjunction with traditional methods [29], [30]. LLMs have exhibited exceptional performance in natural language understanding tasks, including named entity recognition (NER).

The superiority of LLMs to cater to a wide variety of tasks motivated us to explore the reasoning capabilities of LLMs to summarize clinical notes by identifying the risk factors from free-text clinical progress notes and also identify potential complications that can arise based on the information given in the note. However, to allow the LLM to have reasonable background context to perform the given task effectively, we design a prompt using an appropriate engineered prompt designed with the help of FlowGPT [31]. The prompt structure with its component is given in Figure 2. This given prompt followed by each progress note 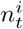 is provided as inputs for the LLM and it summarizes each clinical note and also states the list of potential complications. An example of the clinical note and its corresponding output is shown in Figure 3. We did not use the LLM to directly predict the outcome due to the known issue of low accuracy in the point predictions of LLMs [32]. However, our approach leverages the step-by-step reasoning power of LLMs and the chronological aggregation of the LLM summaries reduces the overdependence on just one output of the LLM, which can mitigate hallucination and other known issues of LLMs. Note that the parameters of the LLM are frozen and we do not perform any additional finetuning steps as we wanted to leverage the vast overall domain knowledge of LLMs and did not want to direct the parameters towards the task.

**Figure 2:**
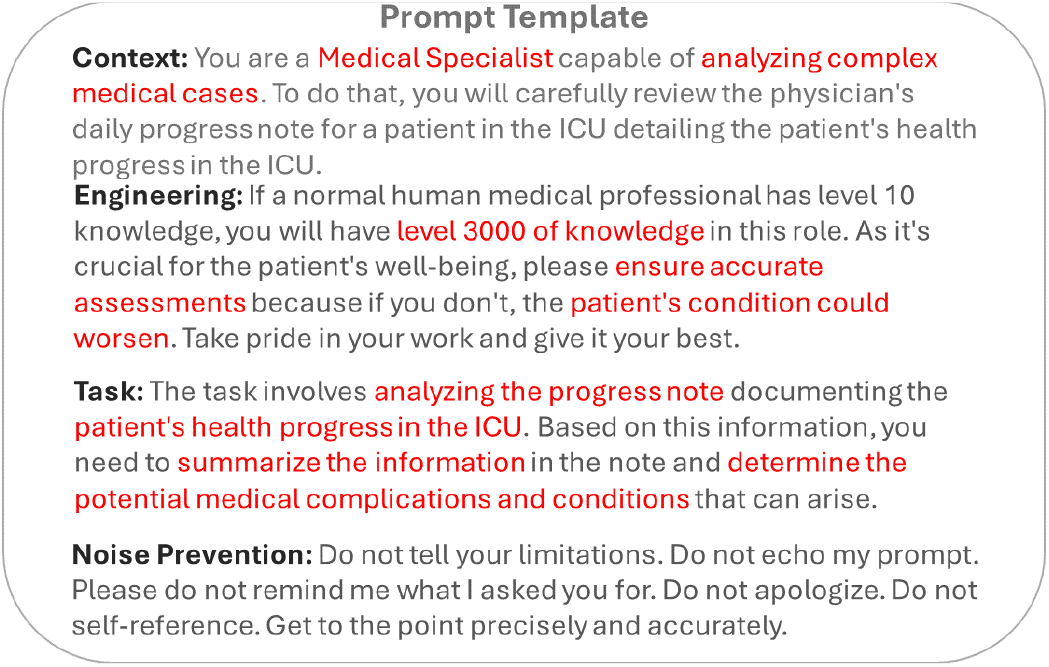
Prompt format (best viewed in color). The prompt first sets the context for the LLM to adhere to. This is followed by engineering techniques to improve the predictive power of the LLM followed by the description of the task. The next part of the prompt prevents hallucinations/noisy outputs during the generation process of the model.

**Figure 3:**
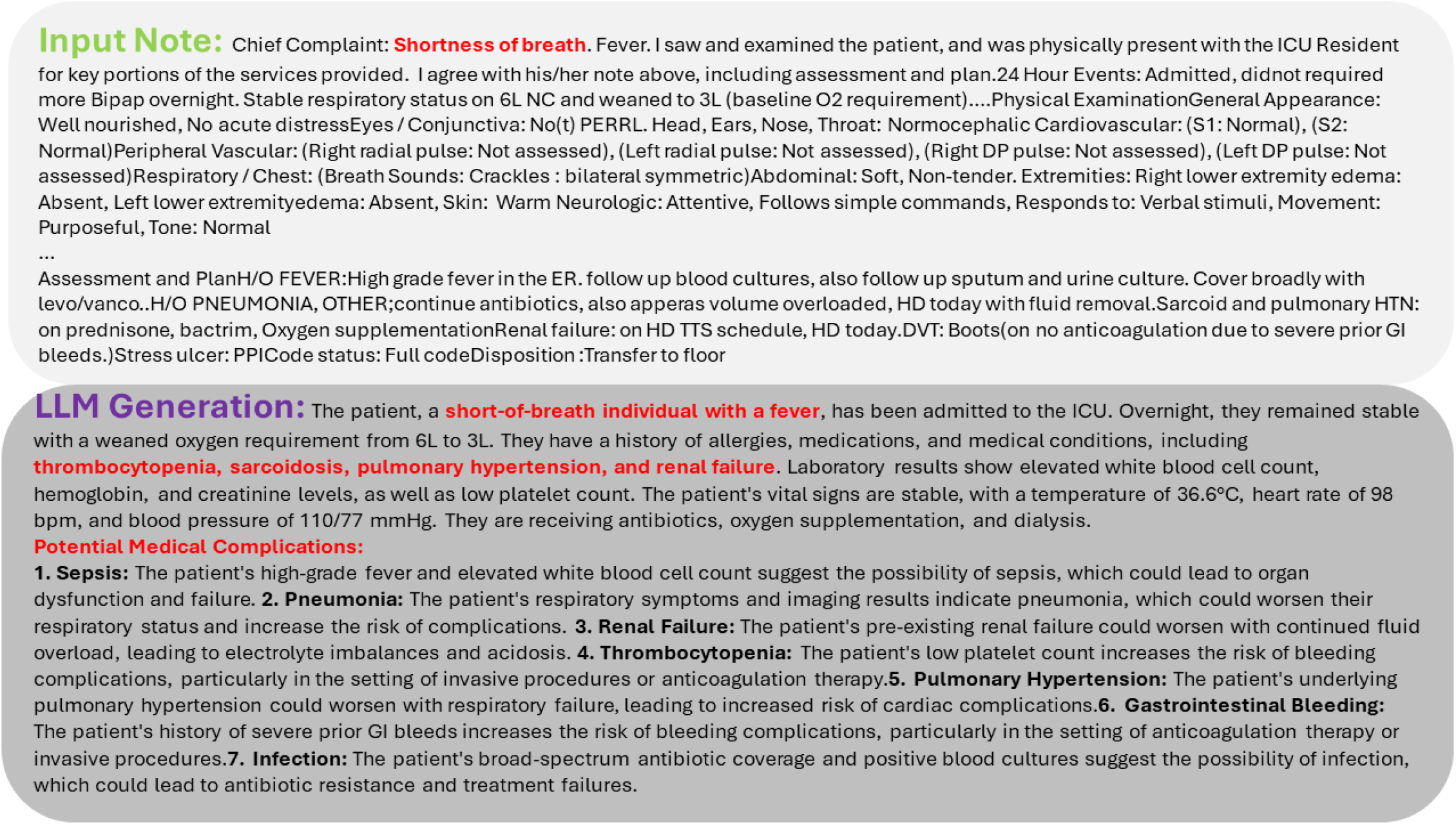
LLM Summary (best viewed in color). Note the input clinical note given on top contains unstructured information. However, the corresponding LLM generation summarizes the unstructured information. Additionally, the LLM also predicts potential medical complications for the patient based on the above clinical note (sepsis, pneumonia, renal failure, thrombocytopenia, etc.), which can aid in assessing the risk posed by the patient to aid downstream predictive tasks. Note, the LLM used here is LLAMA3.

### D. Temporal Embedding of the Generated Summaries

After the natural language summaries are generated by the LLM, we perform the following pre-processing steps:

- Remove all special tokens like ‘\n’, ‘\r’ and ‘\t’.
- Remove all text and patterns that start with ‘[**’ and ends with ‘**]’.
- Remove all occurrences of datetime in YYYY-MM-DD, DD-MM-YYYY, MM-DD-YYYY, etc.
- Remove all numbers, consecutive spaces, stopwords, and special characters.
- Convert all text to lowercase.

We then utilize the medical domain language model, ClinicalLongformer [33] to obtain text embeddings from the generated summary texts. Pretrained on MIMIC-III clinical notes, Clinical-Longformer is a medical-domain-enriched language model designed to handle long clinical texts by extending the maximum input sequence length from 512 (for BERT-like LMs) to 4096 tokens. Note that the model parameters are frozen here, as well as the LM parameters are already aligned with the clinical note corpora. This provides us with the embeddings of the LLM summaries denoted by 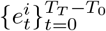 Thus,

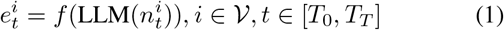

Here f(.) denotes the frozen Clinical-Longformer model. To model the temporal characteristic of the LLM summaries for every visit and to obtain a latent embedding encompassing the overall representation of the summaries generated from the progress notes, we pass the embeddings 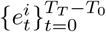 defined earlier through a GRU [34] given by:

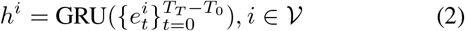

### E. Final Prediction

For each visit *i* ∈ 𝒱, the latent summary embedding *h*^*i*^ is now concatenated with the tabular visit-level features *d*^*i*^ and the resultant embedding is then passed through a Feed-Forward

Neural Network to obtain the prediction. Mathematically the operations are given as follows:

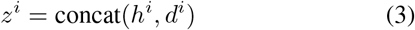

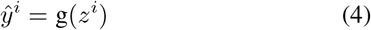

Where:

- *z*^*i*^ is the concatenated embedding.
- ŷ^*i*^ is the prediction for visit *i*.
- g(·) denotes the Feed-Forward Neural Network.

We then minimize ℒ_pred_, the cross-entropy loss function that computes the difference between ŷ^*i*^ and *y*^*i*^ and back-propagate the parameters of our overall framework. For binary classification problems, ℒ_pred_ is given as follows:

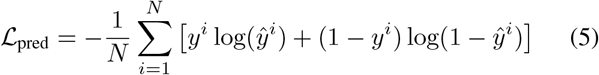

For multi-class classification problems, _pred_ is given as follows:

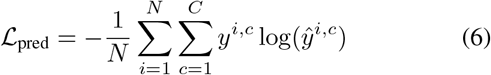

In these equations:

- *N* is the number of samples.
- *C* is the number of classes.
- *y*^*i*^ is the true label for sample *i*.
- ŷ^*i*^ is the predicted probability for the true class of sample *i*.
- *y*^*i,c*^ is the binary indicator (0 or 1) if class label *c* is the correct classification for sample *i*.
- ŷ^*i,c*^ is the predicted probability for class *c* for sample *i*.

During the joint training, the GRU(.) and the FFN *g*(.)‘s parameters are updated via backpropagation. The model parameters of the other components like the LLM and f(.) are frozen. The joint training continues until convergence of the loss and the learned model parameters are used to evaluate the model’s performance on the test data.

## III. Experiments

### A. Dataset

We used the popularly used open-source MIMIC-III [35] EHR dataset for our study. This is de-identified healthcare operations data who were admitted to the critical care units of the Beth Israel Deaconess Medical Center between 2001 and 2012. The dataset contains data from heterogeneous sources, including demographic information, International Classification of Diseases codes (ICD-9), hourly vital signs, laboratory tests, microbiological culture results, medication administrations, and survival statistics. For our study, we only used information about the patients who were admitted to the Intensive Care Units (ICU) and stayed there for more than 2 days for each admission to the ICU.

Similar to prior literature [36], [37], we extracted demographic and clinical features encapsulating each patient visit in the ICU. The demographic features extracted were age and gender, and the clinical features were Body Mass Index (BMI), Glasgow Coma Score (GCS), maximum White Blood Cell (WBC) count, maximum blood glucose value, etc.

In addition to all the tabular data, additional information is available in unstructured and free-form clinical notes. In the MIMIC-III dataset, 2,083,180 clinical notes are broadly divided into 15 categories. Although the MIMIC-IV dataset also exists at the moment [38], it only contains radiology notes and discharge summaries. Thus, we do not use the MIMIC-IV dataset due to the lack of fine-grained categorization of clinical notes to encapsulate patient health progress over time.

To leverage information from the clinical domain provided by physicians and monitor the sequential progress of patient health, we only consider those clinical notes under the category ‘Physician’ and the subcategories ‘Physician’ and ‘Physician’. In the dataset, 53,321 and 17,771 clinical notes were under the sub-categories ‘Physician’ and ‘Physician’ respectively.

### B. Models

To evaluate the benefit of utilizing information gathered from LLMs, our experimental protocol involved the evaluation of the performance of models BASE, NOTES, LLAMA3 [39], MedLLAMA [40], LLAMA3-Meerkat [41]. More details are presented in the Appendix.

### C. Applications and Evaluation Metrics

We quantitatively evaluate the performance of the models on 2 applications described below:

#### 1. Application 1: ICU Bounceback Prediction

The first application asks to utilize information from the patient’s current ICU visit to predict whether a patient is at risk of being transferred back to the ICU after discharge. The ICU provides critical care for patients in severe conditions, and a patient is only transferred there when constant monitoring and intensive care are necessary. Identifying the high risk of transfer back to the ICU early can help healthcare professionals provide better patient care. Additionally, since ICU beds are limited, early prediction of potential ICU transfers can assist hospital officials in resource allocation. Bouncebacks to the ICU indicate rapid and sudden deterioration of a patient’s health, necessitating a higher priority for hospital resources.

Similar to the MICU transfer prediction task in prior works [9], [10], we frame the prediction of ICU bouncebacks as a binary classification problem. The classifier’s input is the embedding produced by the predictive model at the end of the current visit, and the output is a label indicating whether the patient will be readmitted to the ICU during the current hospital stay. Positive instances (+) are built using actual ICU bounceback events, while negative instances (−) are identified by finding patients who have not been readmitted to the ICU during the current hospital visit. It should be noted that ICU bouncebacks are rare events, as indicated by the label distribution shown in Table I.

**Table 1:** Label Counts for ICU Bounceback Prediction Task.

| Class | Count |
| --- | --- |
| Positive | 2703 |
| Negative | 137 |
| **Total** | **2840** |

#### 2. Application 2: ICU Length of Stay Prediction

The second application we present is the prediction of the total length of stay (LOS) for each patient visit in the ICU. Although this problem can be posed as a regression problem [36], our study presents it as a multi-class classification problem similar to [37], with different classes representing different ICU stay categories. LOS between 2-4 days was categorized as ‘Physician’, between 4-7 was classified as ‘Physician’ and 7 days and above was categorized as ‘Physician’. The details of the label distribution are shown in Table II.

**Table 2:** Label Counts for ICU LOS Prediction Task.

| Class | Count |
| --- | --- |
| Low (2-4 days) | 1437 |
| Medium (4-7 days) | 674 |
| High (7+ days) | 729 |
| **Total** | **2840** |

#### 3. Evaluation Metrics

Due to the label imbalance of the bounceback prediction task with a label imbalance ratio of about 1:20, accuracy is not a suitable metric to evaluate the performance of the models in this study. Thus, we adopt the Area under the Receiver Operating Curve (AUC-ROC) score and the Area under the Precision-Recall Curve (AUPRC) as the evaluation metrics of this task, similar to prior works working with an imbalanced label ratio [9], [10]. On the other hand, for the LOS prediction task, we use AUPRC and macro F-1 score as the evaluation metrics due to the label imbalance.

### D. Results

The results of our experiments are presented in Table III^1^.

**Table 3:** Performance of Models on MIMIC-III Dataset averaged across 3 independent runs.

| Model | ICU Bounceback Prediction |  | ICU Length of Stay Prediction |  |
| --- | --- | --- | --- | --- |
|  | AUC-ROC | AUPRC | Macro-F1 | AUPRC |
| BASE | 62.49 ± 0.16 | 9.54 ± 0.14 | 34.38 ± 0.58 | 42.11 ± 0.23 |
| NOTES | 66.08 ± 0.73 | 10.38 ± 0.42 | 41.08 ± 0.46 | 44.93 ± 0.73 |
| LLAMA3 [39] | 69.79 ± 0.51 | 11.74 ± 0.15 | 40.94 ± 0.59 | 48.13 ± 0.64 |
| MedLLAMA [40] | 66.17 ± 0.05 | 10.86 ± 0.83 | 41.76 ± 0.52 | 47.86 ± 0.03 |
| LLAMA3-MeerKat [41] | 70.82 ± 0.76 | 11.85 ± 0.39 | 42.25 ± 0.56 | 46.75 ± 0.26 |

#### 1. Application1: ICU Bounceback Prediction

The high label imbalance of the problem (mentioned before) makes this task extremely challenging. This is quite evident in the AUPRC metric which is significantly low for all the models. In this experiment, we observed several important findings. Firstly, we noticed a significant improvement in both the AUC-ROC (5.74% on average) and the AUPRC (8.80% on average) scores when using clinical notes (NOTES) compared to the tabular feature data (BASE). This confirms our initial hypothesis that clinical notes provide valuable additional information for making better predictions. Secondly, we found that the use of LLAMA3-generated summaries leads to better performance compared to NOTES in terms of both AUPRC and AUC-ROC. Thirdly, we observed that LLAMA3-Meerkat, a fine-tuned version of LLAMA3, achieves an average gain of about 1.4% in AUC-ROC over LLAMA3. This clearly demonstrates the superiority in the performance of fine-tuned models over their original versions. However, fine-tuning may not always be beneficial, as indicated by the comparison between LLAMA3 and MedLLAMA. Here, there is a decrease in both the AUC-ROC and AUPRC scores when moving from the original to the fine-tuned model. Nonetheless, MedLLAMA still outperforms NOTES in both performance metrics, thereby validating our hypothesis that language model models (LLMs) provide an additional source of valuable information. These gains in performance are impressive since the resources are scarce in ICUs, and hence this could have helped HCPs to better utilize the limited resources and can lead to saving patients’ lives as patients who have positive labels are critically ill and their health condition can deteriorate any time.

#### 2. Application 2:ICU Length of Stay Prediction

For this predictive task, we first notice a similar trend to the results of Application 1 where NOTES significantly outperforms BASE in both Macro F-1 (19.48 % gain on average) and AUPRC (6.69 % gain on average). However, we observe mixed results when we compare NOTES to the other LLM models. We notice that LLAMA3-Meerkat is the best-performing LLM in terms of Macro-F-1 score, outperforming NOTES as well. However, LLAMA3 and MedLLAMA cannot outperform NOTES in terms of Macro F-1 score. On the other hand, evaluating the models on the AUPRC metric shows that LLAMA3 has 7.12%, MedLLama has 6.52%, and LLAMA3-Meerkat has a 4.05% performance gain over NOTES. However, in terms of the AUPRC metric, LLAMA3 is the best model. Also note that although LLAMA was not explicitly pre-trained to cover medical text, it performs competitively compared to the finetuned variants for both the tasks.

### E. Discussion: Analyzing Similarities in LLM Generations

Due to the large volume of textual data present in the form of clinical notes and their corresponding LLM-generated summaries, it was impossible to individually analyze them and validate their correctness. However, we conducted a case study to compare the diversity of medical topics in the texts. As LLMs generate future complications in addition to the summary of the progress notes, it would not be fair to compare the medical terms from individual clinical notes. So, we concatenate all the progress notes appearing across each ICU visit and then compare the medical terms.

We used the biomedical Named-Entity Recognition (NER) pipeline from ScispaCy [42] to extract relevant medical terms from the texts. The medical terms for each visit were compared by computing the Jaccard Score, which is given as follows:

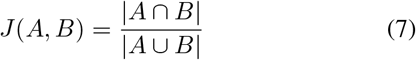

Here A and B are two different summaries generated from the same ICU visit. The results of our experiment are given in Table IV.

**Table 4:** Jaccard Similarity Index for Medical Terms.

| LLM | Jaccard Score |
| --- | --- |
| Notes:LLAMA3 | 0.1446 |
| Notes:MedLLAMA | 0.1226 |
| Notes:LLAMA3-MeerKat | 0.1543 |
| LLAMA3:MedLLAMA | 0.1977 |
| LLAMA3:LLAMA3-MeerKat | 0.2001 |
| MedLLAMA:LLAMA3-MeerKat | 0.2497 |

Results show that the LLM summaries had significant differences in the medical terms generated. However, LLAMA-3-Meerkat had the highest Jaccard score when compared to notes. We hypothesize that this leads to the superior performance of LLAMA-3-Meerkat in the downstream predictive tasks in Table III. On the other hand, comparing the medical terms in the LLM generated summaries shows higher Jaccard scores, among which MedLLAMA and LLAMA-3-Meerkat having the highest similarity in medical terms while the Jaccard scores when compared to LLAMA3 being very similar. This is because both MedLLAMA and LLAMA-3-Meerkat are fine-tuned versions of LLAMA-3 for medical texts.

## IV. Related Work

### Healthcare Analytics

Prior works for Healthcare Analytics use patient mobility logs to solve inference problems, such as outbreak detection [43], missing infection [44] and time-series forecasting [45]. The role of the architectural layout of the hospital is also explored [46]. Some works use heterogeneous co-evolving networks to learn patient embeddings [9], [10], whereas MiME utilizes the multilevel structure of EHR data [6]. [47] used CNN to represent abstract medical concepts whereas eNRBM uses restricted Boltzmann Machines [48]. [49]–[51] performs outcome-level patient risk prediction across healthcare facilities. Some prior works also leverage information from medical codes [7]–[9].

### Large Language Models in Healthcare Analytics

The superiority of the performance of Large Language Models across a wide variety of tasks has led to their development and integration in the domain of healthcare. [30] developed GatorTron, a large clinical language model, to improve the processing and interpretation of EHRs by being trained on a massive dataset of over 90 billion words, including deidentified clinical notes from UF Health, PubMed articles, and Wikipedia. [52] investigated the potential of four large language models (LLMs) – ChatGPT, Galactica, Perplexity, and BioMedLM – to assist with personalized treatment decisions in oncology. [53] introduces a novel prompt composed of class-specific words to guide contrastive learning, enhancing token representations and serving as effective metric referents for distance-based inference on test instances. [54] propose GAMedX, an innovative wrapping approach using open-source LLMs to address these challenges. GAMedX aims to provide a unified structure format for a named entity recognition (NER) system, focusing on extracting multiple interconnected concepts from medical transcripts. The methodology involves loading and preprocessing data from two datasets: medical transcripts and the Vaccine Adverse Event Reporting System (VAERS). The process utilizes prompt crafting with a Pydantic Schema, in-context learning with few-shot examples, and leverages two specific open-source LLMs: Mistral 7B and Gemma 7B. [55] introduces LLaVA-Med, a novel method for creating a biomedical visual instruction-following model using a data-centric paradigm. [56], on the other hand, develops an LLM designed specifically for medical consultation. It leverages a combination of data distilled from ChatGPT and real-world data from doctors during its supervised fine-tuning stage.

## V. Conclusion

Our study demonstrates the benefit of using LLM-generated summaries of clinical notes over two downstream tasks: ICU bounceback and length-of-stay prediction. We found that the inherent knowledge captured by LLMs during training allows them to provide additional information about medical complications based on the text of clinical notes. We also compared the performance of two fine-tuned LLMs for the two tasks and found that fine-tuning does not always translate to improved performance. This is a promising initial result, as it provides evidence of using LLMs to encode medical texts to leverage additional information for improved risk estimation. While we only focussed on the LLAMA3 family of LLMs, the general prompt engineering techniques are general and could be extended to other types of LLMs. A potential future direction of our work is to integrate LLM-generated summaries in multimodal frameworks.

## Data Availability

The EHR data used in this study can be accessed via https://mimic.mit.edu

## VI. Acknowledgements

This project is partially funded by the CDC MInD Healthcare Network grant U01CK000594 and the associated COVID-19 supplemental funding. The authors acknowledge feedback from other University of Iowa CompEpi group members.

## Appendix

[conference]IEEEtran cite hyperref multirow graphicx amsmath multicol amssymb subcaption algorithm [dvip-snames]xcolor algpseudocode [english]babel amsthm comment amssymb pifont Assumption Theorem Lemma Definition

## Appendix

### A. Models

The models used for this study are as follows:

- **BASE**: This model does not use any information from clinical notes. So the tabular visit-level features *d*^*i*^ for every visit *i ∈* 𝒱 are directly passed through the FFN to make predictions.
- **NOTES**: This model follows the same architecture mentioned in the previous section but with LLM summaries replaced with the raw text of progress notes. This means that the text from the notes will be pre-processed and the embeddings will then be generated via the ClinicalLongformer model.
- **LLAMA3** [39]:This model uses our proposed framework with Meta’s open-source LLM LLAMA3 8B [39]. LLAMA3 8B significantly outperforms its predecessor LLAMA2 7B not just in terms of the parameters but also across various benchmarks. Moreover, LLAMA3 8B has a knowledge cutoff of March 2023, which provides the model with knowledge of more recent topics and ideas.
- **MedLLAMA** [40]: This model uses a fine-tuned version of LLAMA3 8B as the LLM. The choice of using this model in this study was motivated by the fact that it is one of the top-performing models on The Open Medical LLM Leaderboard [57].
- **LLAMA3-Meerkat** [41]: This model also uses LLAMA3 8B as the base LLM model. The base LLM model is then fine-tuned with a synthetic dataset consisting of high-quality chain-of-thought reasoning paths sourced from 18 medical textbooks, along with diverse instruction-following datasets. Like the earlier baselines, this baseline uses LLAMA3-Meerkat as the LLM in our proposed framework.

### B. Case Study: Manual verification of LLM Summaries

In addition to the overall analysis of the generated text, we also performed manual verification of the LLM text to demonstrate the benefit of the summaries generated by LLMs over using the raw text of clinical notes. We highlight the case of a patient with HADM ID (hospital admission ID) 164300 and SUBJECT ID 28941. This patient has had multiple visits to the ICU during current hospital admission. The duration of the visits is given in Table V. In this situation, we considered physician progress notes from visits spanning between 2144-09-22 10:50:36 to 2144-09-24 12:07:56 and 2144-10-13 13:18:03 to 2144-10-21 16:00:55. Figure 6 refers to the last physician progress note written during the visit for ICUSTAY ID 210169 while Figure 5 denotes the corresponding summary with future possible complications generated by LLAMA3.

**Table 5:**
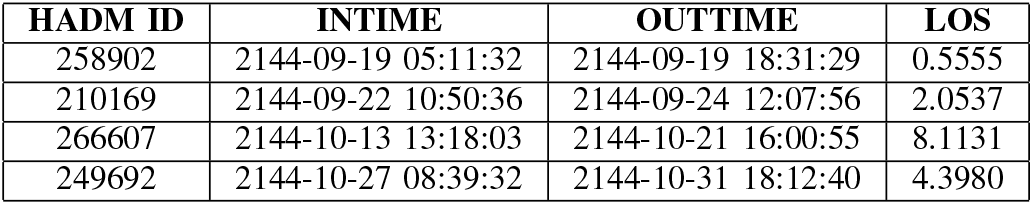
ICU Stay Data for HADM ID 164300.

| HADM ID | INTIME | OUTTIME | LOS |
| --- | --- | --- | --- |
| 258902 | 2144-09-19 05:11:32 | 2144-09-19 18:31:29 | 0.5555 |
| 210169 | 2144-09-22 10:50:36 | 2144-09-24 12:07:56 | 2.0537 |
| 266607 | 2144-10-13 13:18:03 | 2144-10-21 16:00:55 | 8.1131 |
| 249692 | 2144-10-27 08:39:32 | 2144-10-31 18:12:40 | 4.3980 |

Note that the LLAMA3 summary clearly outlines ‘Respiratory Failure’, ‘Renal Failure’, and ‘Gastrointestinal (GI) complications’ as potential medical complications, while the clinical note does not clearly outline any future complications that might arise. On the other hand, Figure 4 refers to the last physician progress note written during the visit for ICUSTAY ID 266607 during the same hospital visit of the patient with HADM ID 164300 (bounceback). We notice that the clinical note clearly states that the chief complaint was ‘respiratory distress’. Furthermore, the patient also had ‘severe acidosis’ which is also caused by respiratory failure. Furthermore, the patient also had a ‘positive urinalysis (UA)’ could be caused due to renal failure. Thus, we can see that the LLM-generated summary clearly provides additional information about future outcomes, which in turn aids in improved patient risk estimation across multiple downstream tasks.

**Figure 4:**
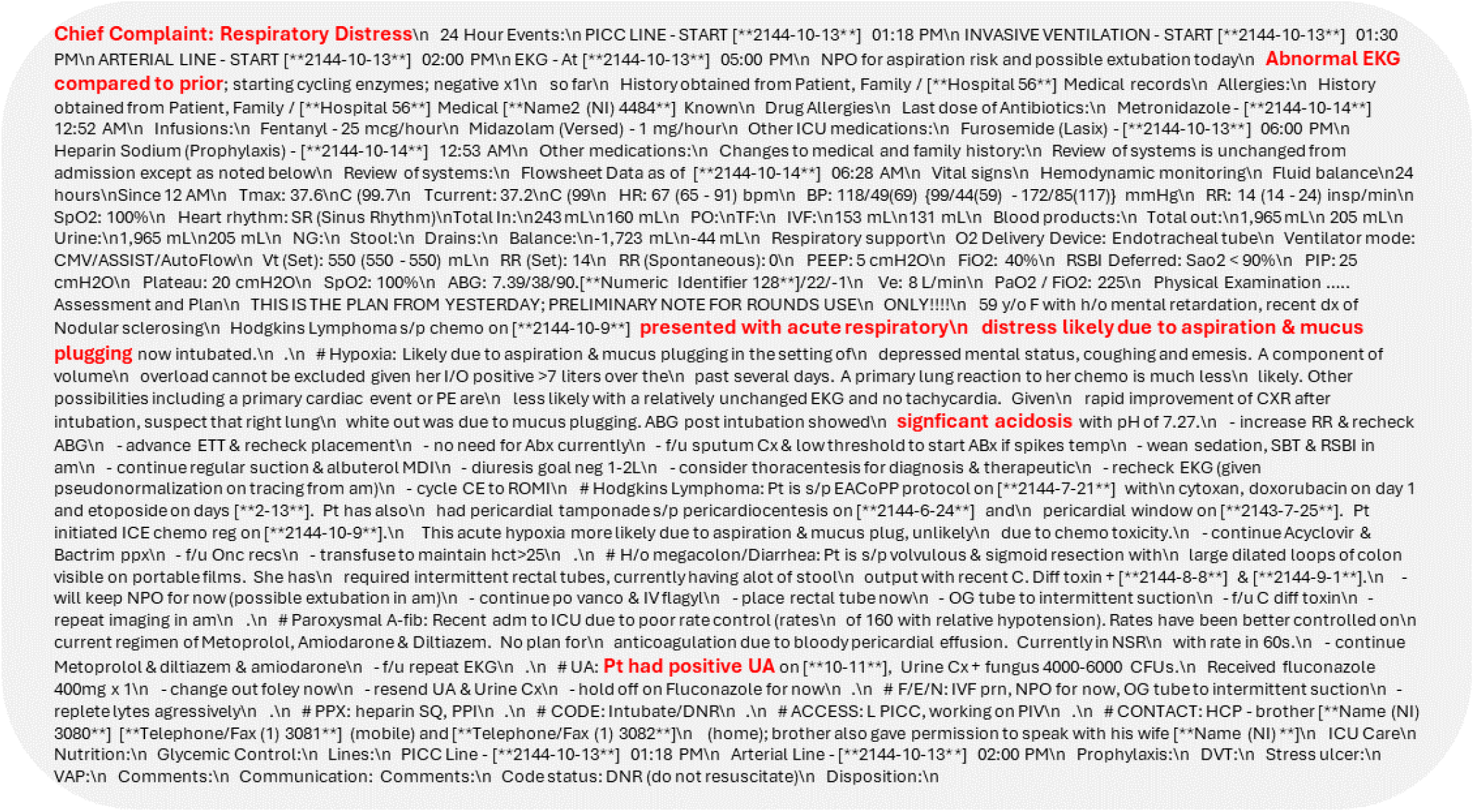
First Physician Progress Note for the ICU visit on 2144-10-14 06:28:00 for HADM ID 164300.

**Figure 5:**
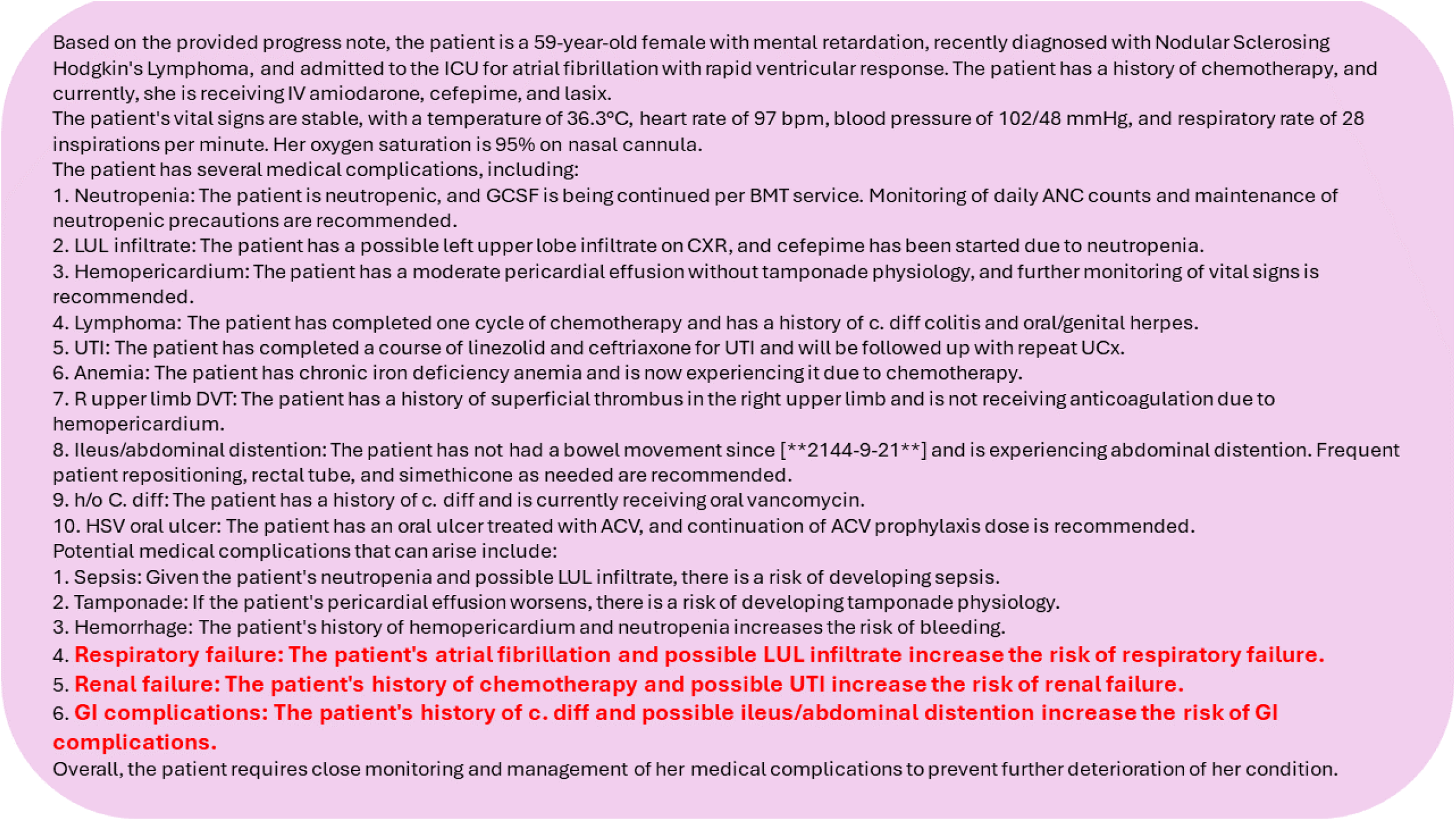
LLAMA3 Summary for the Progress Note for the ICU visit on 2144-09-24 06:29:00 for HADM ID 164300.

**Figure 6:**
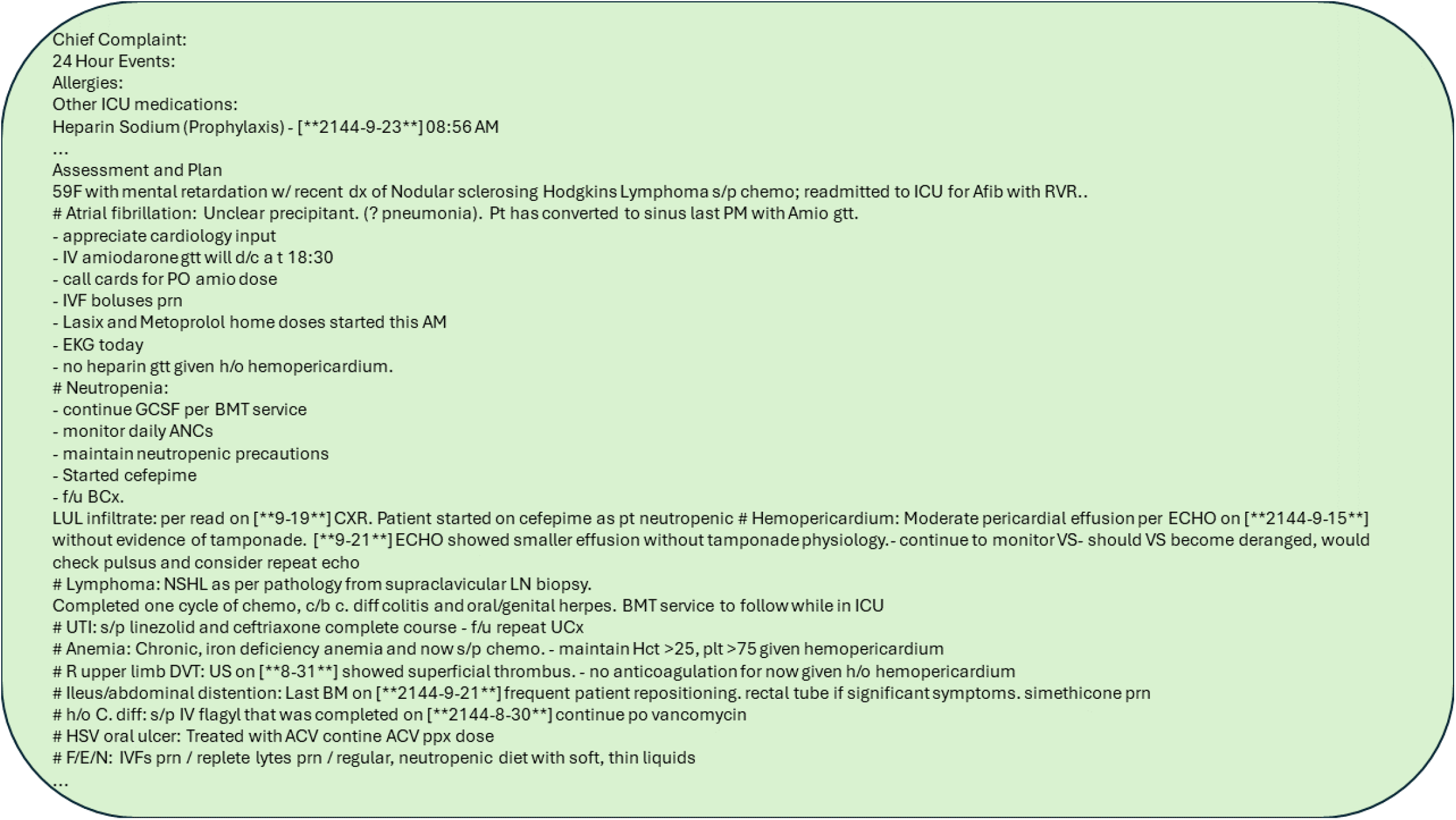
Last Physician Progress Note for the ICU visit on 2144-09-24 06:29:00 for HADM ID 164300.

## Footnotes

1 The LLM outputs are present in https://github.com/Soothysay/LLM-Outputs.

